# Learning Health System for implementation, scale-up, and sustainment: A systematic review to consolidate guidance for improvement

**DOI:** 10.1101/2025.09.24.25336590

**Authors:** Cassandra Lane, Sam McCrabb, Heidi Turon, Caitlin Bialek, Lucy Couper, Magdalena Wilczynska, Samantha Gray, Courtney Barnes, Madeleine Fee, Tanja Kuchenmüller, Davi Mamblona Marques Romao, Luke Wolfenden

**Affiliations:** School of Medicine and Public Health, The University of Newcastle, Newcastle NSW; Australia; Hunter New England Population Health, Hunter New England Area Health Service, Newcastle NSW; Australia; National Centre of Implementation Science, The University of Newcastle, Newcastle NSW; Australia; Hunter Medical Research Institute, New Lambton Heights NSW; Australia; Research for Health Science Division, World Health Organization

**Keywords:** Learning Health System, Implementation, Scale-up, Sustainment, Evidence Synthesis, Framework Synthesis

## Abstract

**Background:** Learning Health Systems (LHSs) enhance the integration of research and health service delivery by generating timely, contextually relevant evidence to guide decision-making and continuous improvement. Although various LHS frameworks exist, practical guidance for operationalising LHSs to support the implementation of health interventions remains limited. This systematic review aimed to consolidate existing guidance to identify the supportive infrastructure (pillars) and improvement processes (steps) required to improve the implementation (including scale up or sustainment) of health programs, policies, or practices.

**Methods:** We systematically searched five academic databases and grey literature sources for documents describing LHSs, as well as process models, guidelines, or tools for improving implementation, scale-up, or sustainment of health interventions. Title, abstract, and full-text screening were conducted independently by two reviewers. Data were synthesised separately for pillars and steps. Framework synthesis was used as the first analytic stage to identify pillars and steps, starting with a codebook informed by an existing LHS framework and refined iteratively throughout the coding process. Thematic Synthesis was then used to identify patterns and concepts within each pillar and step.

**Findings:** From 12,151 records and 25 websites, 96 unique guidance documents were included. Six Pillars were identified as important to operationalise LHS improvement processes: 1-Interest holder engagement, 2-Workforce development and capacity, 3-Evidence surveillance and synthesis, 4-Data collection and management, 5-Governance and organisational processes, and 6-Cross-cutting infrastructure. The improvement process was comprised of 10 ‘Steps’ across three LHS phases: Phase 1) Knowledge to Practice -Identify and understand the problem; Decide and plan for action; Assess and build capacity; Pilot; Phase 2) Practice to Data - Execute the action; Collect data; Monitor and respond; Phase 3) Data to Knowledge-Analyse and evaluate; Disseminate; and Decide (continue, adapt, or cease improvement efforts). Despite the diversity in purpose and context across included documents, the consolidated steps and pillars were conceptually consistent, suggesting a shared foundation. Some contextual variation in emphasis and operationalisation was noted, particularly among guidance focused on scale-up or sustainment.

**Conclusions:** This review synthesised diverse guidance to consolidate LHS pillars and improvement steps to better implement, scale or sustain health interventions. The findings offer a structured yet adaptable approach for operationalising LHSs focused on implementation, scale-up, and sustainment. These findings will inform forthcoming WHO guidance and support efforts to strengthen health systems through more systematic and responsive use of evidence.

## INTRODUCTION

Interventions to alleviate much of the health burdens experienced by health systems and communities exist^1,2^. Key impediments to its rapid translation into routine practice include the slow, fragmented processes of transferring evidence from ‘researchers’ to ‘end-users’^3^ as well as a persistent lack of contextually relevant, actionable evidence to inform ‘how to’ best implement and sustain interventions in real-world settings^4^. Addressing these issues requires a fundamental shift in how evidence is produced and applied to enable timely responses to current and emerging health challenges.

International reviews aimed at strengthening health systems and improving research translation have called for better integration of the research and health system enterprises ^5–7^. In Australia, the vision of the landmark McKeon review^7^, for example, was one where research is *“…fully embedded in all aspects of healthcare to deliver: ‘Better Health Through Research’ and achieve the aspiration for Australia to build and maintain the world’s best and most efficient health system”.* It recommends - like those undertaken prior by the United States (US) National Institutes of Health^6^ and the United Kingdom’s Office for Strategic Co-ordination of Health Research^5^ - a reorientation of health systems to better generate and apply health research to improve the quality and outcomes through targeted funding, workforce development, investment in critical research infrastructure, and other strategies.

Learning Health Systems (LHSs) provide a framework by which this could be achieved. LHSs were initially proposed as a way of improving health system performance and impact by the US National Institute of Medicine in 2007^8^. They are defined as systems where “…*science, informatics, incentives, and culture are aligned for continuous improvement and innovation, with best practices seamlessly embedded in the delivery process and new knowledge captured as an integral by-product of the delivery experience”*^9^. In an LHS, health services generate the evidence they need to address their own priorities, with the knowledge created immediately available for decision-making to continuously improve health services and their impact.

Interest in LHSs as a vehicle for improvement is growing. Publications on LHSs have increased markedly over the past decade^10^, and governments made significant investments in infrastructure to reorient health systems toward models of health care aligned with LHS principles. This momentum has been accompanied by the emergence of numerous frameworks and conceptual models of LHSs. While these frameworks vary, they consistently emphasise the importance of foundational ‘pillars’ – such as supportive infrastructure, capabilities, culture and leadership - and continuous, cyclical improvement processes that leverage these pillars to drive data-informed healthcare^10–14^.

Nonetheless, fully operational LHSs remain relatively rare, and their establishment continues to present a considerable challenge. Despite the existence of a range of LHS frameworks, there have been calls for more practical, operational guidance to support their development and functionality.^15^ Given that health systems and services are ultimately responsible for the implementation of health policies, programs, and practices, LHSs offer a promising approach to generating and applying contextually relevant evidence on how best they can do so. Indeed, they offer a potentially transformative means of advancing both implementation science and strengthening health systems and improving patient and population health outcomes^4^. In this context, we conducted a review to inform the development of World Health Organization (WHO) guidance to support the adoption of implementation focused LHSs. This work complements ongoing efforts by WHO and its Evidence-informed Policy Network (EVIPNet) to assist Member States in promoting the systematic and transparent use of evidence in policy^16^. In this review, we focus on synthesis and consolidation of existing LHS frameworks, and models and guidance documents that aim to generate and apply evidence to improve the implementation, and/or scale-up, and/or sustainment of health interventions (policy, practice, program).

Our specific aims were:

1. Pillars Objective: (A) to identify the infrastructure, expertise, and resources (pillars) recommended to support and enable implementation focussed improvement processes – (e.g., ‘steps’); and (B) to describe the core elements or functions within identified pillars.
2. Steps Objective: (A) to describe the steps or stages recommended to improve the implementation (and/or scale-up or sustainment) of health programs, policies, or practice; and (B) to characterise the core actions or activities associated with each identified step.

## METHODS

### Protocol and registration

This review was conducted in accordance with Cochrane best practice methods^17^ and reported following the Preferred Reporting Items for Systematic Reviews and Meta-Analyses (PRISMA) 2020 guidelines^18^. The review protocol was prospectively registered on Open Science Framework (doi: 10.17605/OSF.IO/V4JRC). The protocol was developed by a team of Australia-based implementation and behavioural science researchers, with input of the WHO and a WHO convened Editorial Board of international experts established to support the development of evidence-based guidance.

### Eligibility criteria

Table 1 details the review eligibility criteria.

**Table 1.**
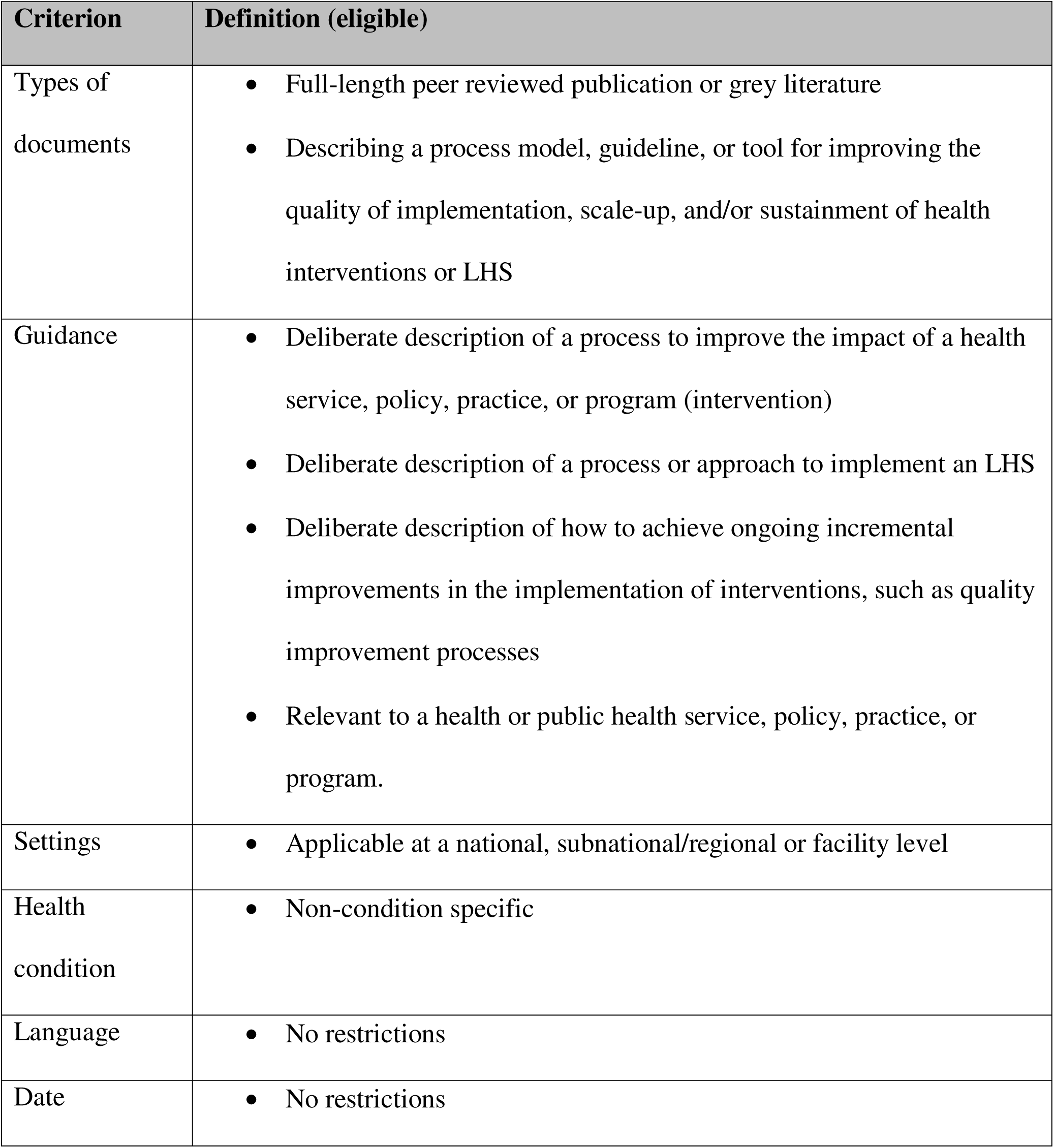
Eligibility criteria.

#### Types of documents

We included documents that described an LHS process, or process model, guideline, or tool (i.e., guidance) intended to improve the quality of implementation, scale-up, and/or sustainment of health interventions. Guidance was defined as a structured and deliberate description of a process to enhance the impact of a health service, policy, practice, or program – typically presented in a series of steps or stages that may or may not be linear^19,20^. Full length documents – defined as those containing substantive narrative content^21^– were eligible for inclusion. Abstracts, conference proceedings, and letters were excluded. There were no language restrictions; articles published in languages other than English, French, Portuguese, Polish, or Spanish (spoken by members of the research team) were translated using Google Translate.

#### Types of guidance

Guidance documents were included if they intended to: i) improve the extent, quality, scale or completeness of the implementation of an intervention or LHS, or ii) sustain such improvements. This included guidance to achieve ongoing incremental improvements in the implementation of interventions (e.g., quality improvement processes) and guidance aimed at supporting implementation at scale. Guidance must have been relevant to a health or public health service, policy, practice, or program. Documents were excluded if they (i) focused solely on a specific condition (e.g., diabetes management plans), (ii) were developed for use outside of health or public health systems (e.g., in manufacturing or agriculture), or (iii) described implementation support activities without a structured process (e.g., training or communication of research findings).

### Search methods for identification of studies

We searched for publications from the last 10 years, although older studies were included if eligible and identified through backward citation searching (described below).

#### Electronic searches

We searched the academic databases MEDLINE (Ovid), PsycINFO (Ovid), and EMBASE (Ovid) in April 2024. Search terms were informed by terminology used in Doherty et al. for implementation research^22^ and by terms related to LHSs^23^ from the Canadian Health Libraries Association identified on the InterTASC Information Specialists’ Sub-Group Search Filter Resource^24^. A research design filter was applied to exclude citations of research trials^25^. The full search strategy is detailed in Supplementary File 1. Eligible documents were also drawn from a recent systematic review and consolidation of adaptation frameworks^21^, as well as any adaptation frameworks published since that review.

#### Searching other resources

In consultation with field experts, we developed a list of key websites to search, including departments within the WHO and other relevant agencies that develop or disseminate guidance to support the implementation or improvement of health services at local, national, or global levels (Supplementary File 2).

#### Backwards citation searching

If an eligible document cited a more complete version of the guidance, efforts were made to locate and include the more comprehensive document in place of the less detailed source.

### Data collection and analysis

#### Selection of studies

After removing duplicates using Endnote and Covidence, title and abstract screening was undertaken independently by two reviewers (SMc, HT, LC, MW, CBi, SG, CBa, KB, MD, BM, RH, NN) using Covidence systematic review software^26^. Full-text articles were retrieved for all publications deemed potentially eligible. Two members of the research team independently assessed the full texts for inclusion and extracted information from the eligible studies (SMc, HT, LC, MW, LW, CBa, KB, MD). Disagreements regarding eligibility were resolved through discussion and, when necessary, consultation with a third reviewer.

#### Data extraction and management

Data from all included studies were extracted by one review author and checked for accuracy by a second reviewer (SMc, HT, LC, MD, MF, MW, SG, KB, CBa). Any discrepancies were resolved through consensus or consultation with a third reviewer (SMc, HT, LC, MD, MF, MW, SG, KB, CBa). A bespoke data extraction form was developed and piloted by two reviewers to ensure consistency and usability. Data were extracted into two broad categories:

- Characteristics of guidance:

- Citation information including name, details, year published.
- Key attributes including intended World Income Level of the country the guidance originated in, stated purpose, intended audience, the discipline in which it originated, and at what level implementation is occurring.
- Details of the steps/phases including the number of steps, the listed steps and definitions, whether the process included a defined endpoint or was continuous, and whether the process was linear or non-linear.

#### Data synthesis

Synthesis methods were informed by the RETREAT criteria for selecting a qualitative evidence synthesis approach^27^ and aligned with the objectives of this review. NVivo software (v14)^28^ was used for data management, coding, syntheses, and sensitivity analyses. Separate synthesis processes were undertaken for the Pillars and Steps objectives by two coding teams, under the direction of an experienced qualitative researcher (CL). To provide an LHS foundation, the codebooks for syntheses were structured based on the LHS Framework by Wolfenden et al^29^ – which was developed based on a review of LHS frameworks by Menear et al^12^ and recommendations for LHSs focused on implementation.

##### Pillars

Framework synthesis^30–32^ was used to identify the infrastructure, expertise, and resources (pillars) recommended to support and enable implementation focussed improvement processes (Objective 1A). The foundation of Framework Synthesis is a strong starting codebook to guide the coding process. The six pillar domains from Wolfenden et al.’s LHS framework^22^ provided the preliminary codebook. All identified guidance documents were coded, starting with the preliminary codebook which was then iteratively developed. A four-person coding team (MW, SG, CL, LW) extracted content including text, table extracts, and figures, though preference was given to narrative content when content was duplicated across formats. Ten frameworks were initially double-coded to refine the codebook and support inter-coder consistency. Remaining frameworks were single-coded and independently checked by a second coder. Discrepancies were resolved through discussion or escalated to a third coder if needed. The coding team met regularly to review and iteratively refine the codebook. A final review and data cleaning were conducted by the synthesis lead (CL).

Thematic synthesis^33^ was then used to describe the core elements or functions within each identified pillar (Objective 1B). The synthesis lead (CL) analysed the coded content within each Pillar to inductively develop themes reflecting specific elements or functions. Original guidance document sources were revisited for clarification where needed, and final themes – presented as narrative summaries within each Pillar – were discussed and confirmed through team consensus.

##### Steps

Framework synthesis^30–32^ was used to synthesise the steps or stages suggested by eligible guidance documents to improve the implementation of evidence-informed health programs, policies, or practice (Objective 2A). The three phases of the LHS Framework by Wolfenden et al^29^ (Phase 1) Knowledge to Practice; Phase 2) Practice to Data; and Phase 3) Data to Knowledge) formed the codebook. Because the phases are quite broad, additional structure was added to the starting codebook. One team member (SMc) first charted the steps described in the first 50 eligible guidance documents in Excel, mapping them against the LHS phases and identifying broad step categories. This initial mapping was checked by another team member (HT) and the output of this process formed the preliminary codebook. A team of four coders (SMc, HT, LC, CB) then followed a similar process to that used for Pillars synthesis: an initial ten guidance documents were double-coded, followed by single-coding with independent checks. The team met regularly to iteratively adapt the codebook, and the synthesis lead (CL) completed final checks and data cleaning.

Thematic Synthesis^33^ was then used to characterise the core actions or activities described within each finalised step (Objective 2B). As with Pillars, this was undertaken by the synthesis lead (CL) and involved inductive thematic coding of the content assigned to each finalised step to identify themes. Guidance documents were revisited where necessary for clarity, and the themes were finalised through team consensus. These discussions also prompted a deductive search for actions or activities that are commonly associated with each step but may not have emerged through the inductive process. This served as a quality assurance check to ensure a more complete and robust synthesis.

#### Sensitivity analyses (Coding comparison)

NVivo software was used to compare coding for Steps and Pillars based on the key attributes extracted (see Data Extraction section).

#### Quantitative coding comparison

NVivo’s cross-tab query function was used to calculate the proportion of frameworks, grouped by key attributes, that contributed to each Step or Pillar. Three members of the team (CL, SMc, HT) visually inspected how much each subtype of a key attribute contributed to the coding of Steps and Pillars, exploring whether any subtypes disproportionately influenced certain Steps or Pillars. For example, whether frameworks from low and middle income countries were under-or over-represented in coding compared with high income countries. This served as a quality check for relative representation across types of frameworks.

#### Qualitative coding comparison

NVivo framework matrices were used to explore the qualitative content coded to each Step or Pillar by framework for the following attributes of interest: World Income Level of the country in which the guidance originated (low and middle income country versus high income country); Level of implementation (national or state policy, regulation, or law versus local or organisational policy, program, guideline); and purpose of the framework (implementation versus scale-up versus sustainability). This thematic analysis, by the synthesis lead, allowed identification of where steps or pillars may need to be operationalised differently according to different attributes. These analyses explored the question: does an LHS pillar or step have different functions or activities across countries of different income levels (high vs low and middle income); when undertaken at different levels of implementation (e.g., national versus local); or used for a different purpose (e.g., implementation versus sustainment)?

## RESULTS

Our search identified 12,151 unique documents and 25 websites. Following screening, 168 documents describing 96 guidance processes, models, or tools were included in this review. **Supplementary file 3** provides details of each guidance document, including label, characteristics, and complete citation. Figure 1 displays the PRISMA flow diagram.

**Figure 1.**
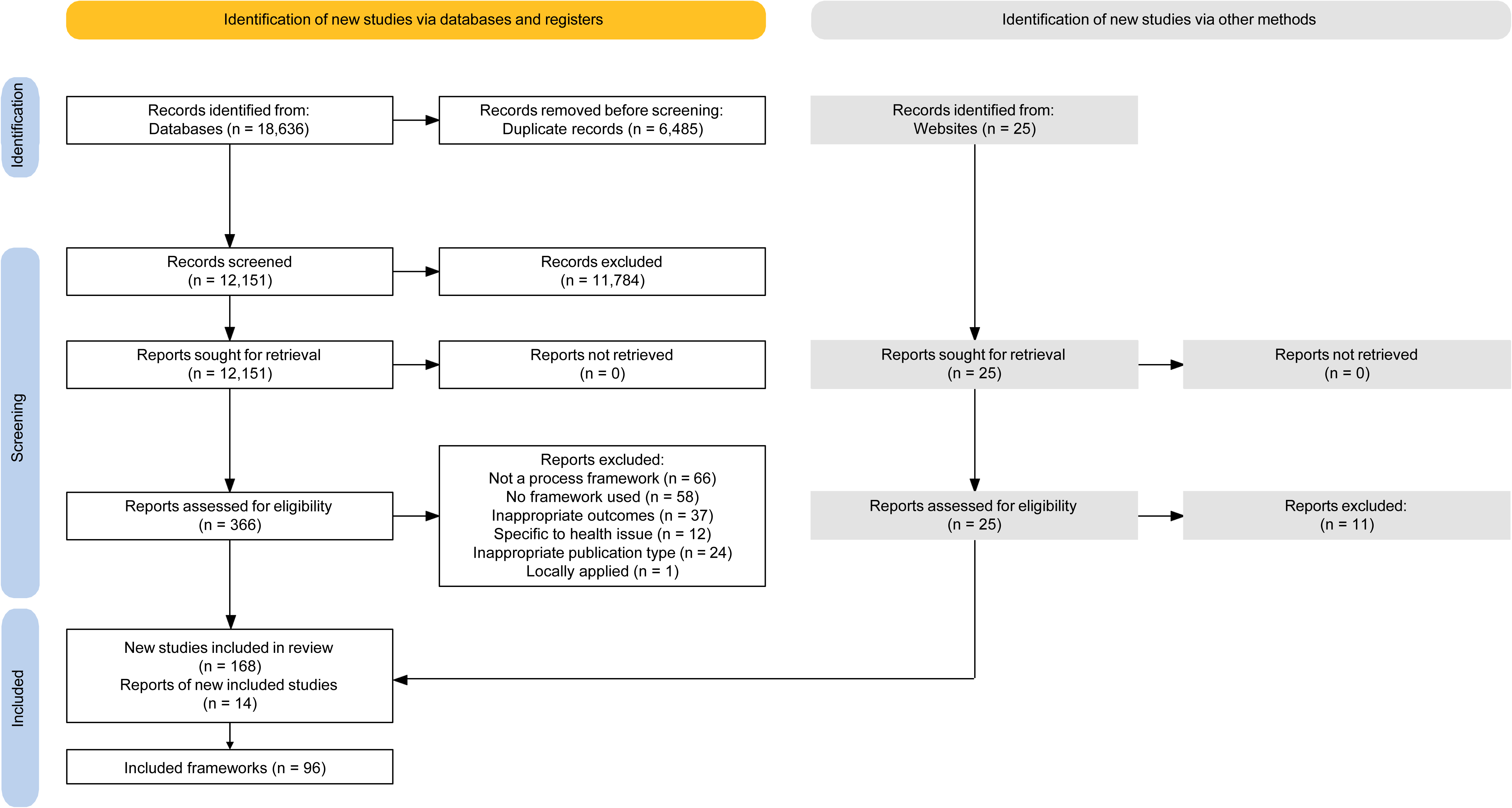
PRISMA flow diagram.

### Characteristics of included frameworks

Most included documents (n=81; 84%) originated from high income country contexts, while a smaller number were applied in low- and middle-income country contexts (n=7; 7%) or in both (n=7; 7%). One (1%) did not include any information to determine the context in which it was applied. The majority were developed within the disciplines of health and public health (n=66; 69%), and the primary purpose for most guidance was implementation (n=68; 71%). Intended end users were predominantly clinicians (n=45; 47%) or multidisciplinary professionals (n=32; 33%). The number of steps outlined in each framework ranged from three to 24, with a mode of five and a median of six steps. Most frameworks were designed for implementation at the organisational policy level (n=63; 66%), while 22 (23%) were applied at both the national/state and local/organisational policy levels and the remaining 11 (11%) at the national/state level. Summary characteristics of guidance documents are detailed in Table 2.

**Table 2.** Summary characteristics of included guidance documents.

| CHARACTERISTIC | DETAILS |
| --- | --- |
| Year published | Range: 1947–2024<br>Mode: 2024 |
| Number of steps | Range: 2–24<br>Mode: 5 |
| Applied in low- and middle-income country | Yes: 16 (17%)<br>No: 80 (83%) |
| Country income level | High-income country: 81 (84%)<br>Low- and middle-income country: 7 (7%)<br>Both: 7 (7%)<br>No information: 1 (1%) |
| Purpose of guidance | Implementation: 68 (71%) |
|  | Scale-up: 7 (7%)<br>Sustainment: 3 (3%)<br>LHS: 11 (12%)<br>Adaptation: (2%)<br>Equity: (2%)<br>De-adoption/implementation: (2%)<br>Multiples: (2%) |
| Intended audience | Clinicians: 45 (47%)<br>Multiple: 32 (33%)<br>Governments/policymakers: 6 (6%)<br>Public health practitioners: 5 (5%)<br>Academics: 3 (3%)<br>Businesspeople: 3 (3%)<br>School teachers: 1 (1%)<br>Implementation support practitioners: 1 (1%) |
| Discipline of origin | Learning Health Systems: 7 (7%)<br>Implementation Science: 15 (16%)<br>Health & Public Health: 66 (69%)<br>Business/Engineering: 6 (6%)<br>Unclear: 2 (2%) |
| Level of implementation | National or State policy, regulation, law: 11 (11%)<br>Local or organisational policy, program, guideline: 63 (66%)<br>Both: 22 (23%) |

### Objective 1: Pillars

Figure 2 displays the resulting LHS for implementation, scale-up, and sustainment. The pillars data synthesis identified six Pillars used to support and enable improvement processes. The coding presence for each Pillar across all guidance documents can be found in Supplementary File 4.

Pillar 1 (Interest holder engagement) are the strategies and supports used involve a diversity of people and organisations with the necessary knowledge and skills, expertise and experience needed for LHS activities. This pillar encompasses consultation, and collaboration and partnerships. Pillar 2 (Workforce development and capacity) are the systems and structures necessary to build, strengthen, and sustain the capacity and capability of personnel involved in operationalising LHSs, including activities related to its pillars and improvement processes (cycles). This might involve mentoring schemes, training mechanisms, and human resource staffing initiatives. Pillar 3 (Evidence surveillance and synthesis) are the systems, structures, and processes used to identify, monitor, and integrate relevant evidence, that may have been generated outside of the LHS, with that generated within it to inform decision-making. This includes infrastructure needed to search, manage, and synthesise different forms of research from multiple and varied sources. Pillar 4 (Data collection and management) are the systems and methods used to capture, process, and apply primary data, and includes (routine) data collection, management and analysis infrastructure. Pillar 5 (Governance and organisational processes) are the structures, standards, culture, and leadership needed for the effective stewardship, operationalisation and coordination of LHS activities. This includes formal reporting, accountability and data governance processes, and regulations and standards to ensure safe and ethical conduct and manage risk. Pillar 6 (Cross-cutting infrastructure) extends across multiple Pillars, encompassing Communication systems (information flow into, within, and out of an organisation); People, expertise and learning (the people involved in operationalisation); Funding and resources (financial, material, and structural supports); and Tools (specific instruments or resources). Table 3 provides an overview of each Pillar and details of its core elements.

**Figure 2.**
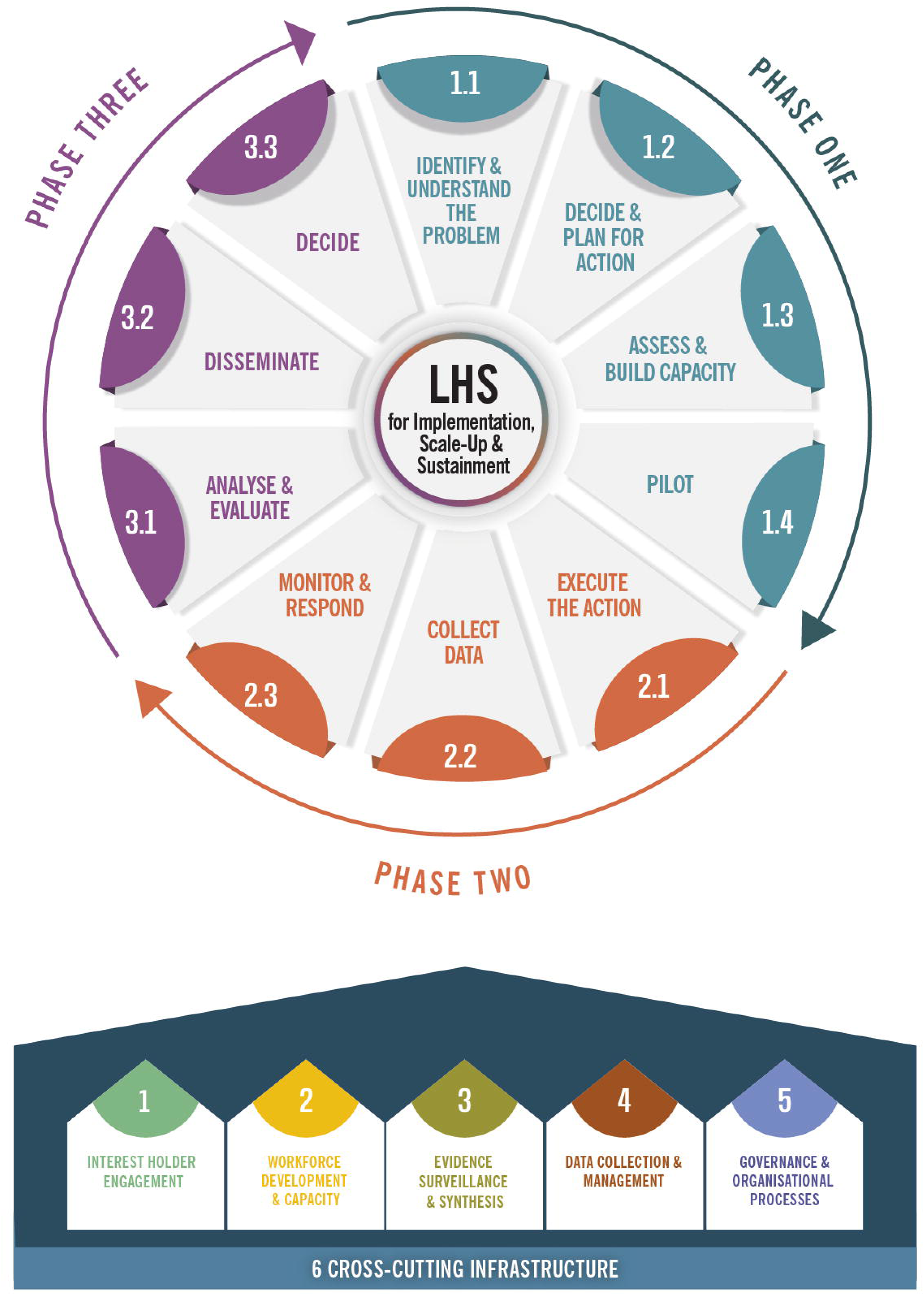
Learning Health System for implementation, scale-up, and sustainment.

**Table 3.**
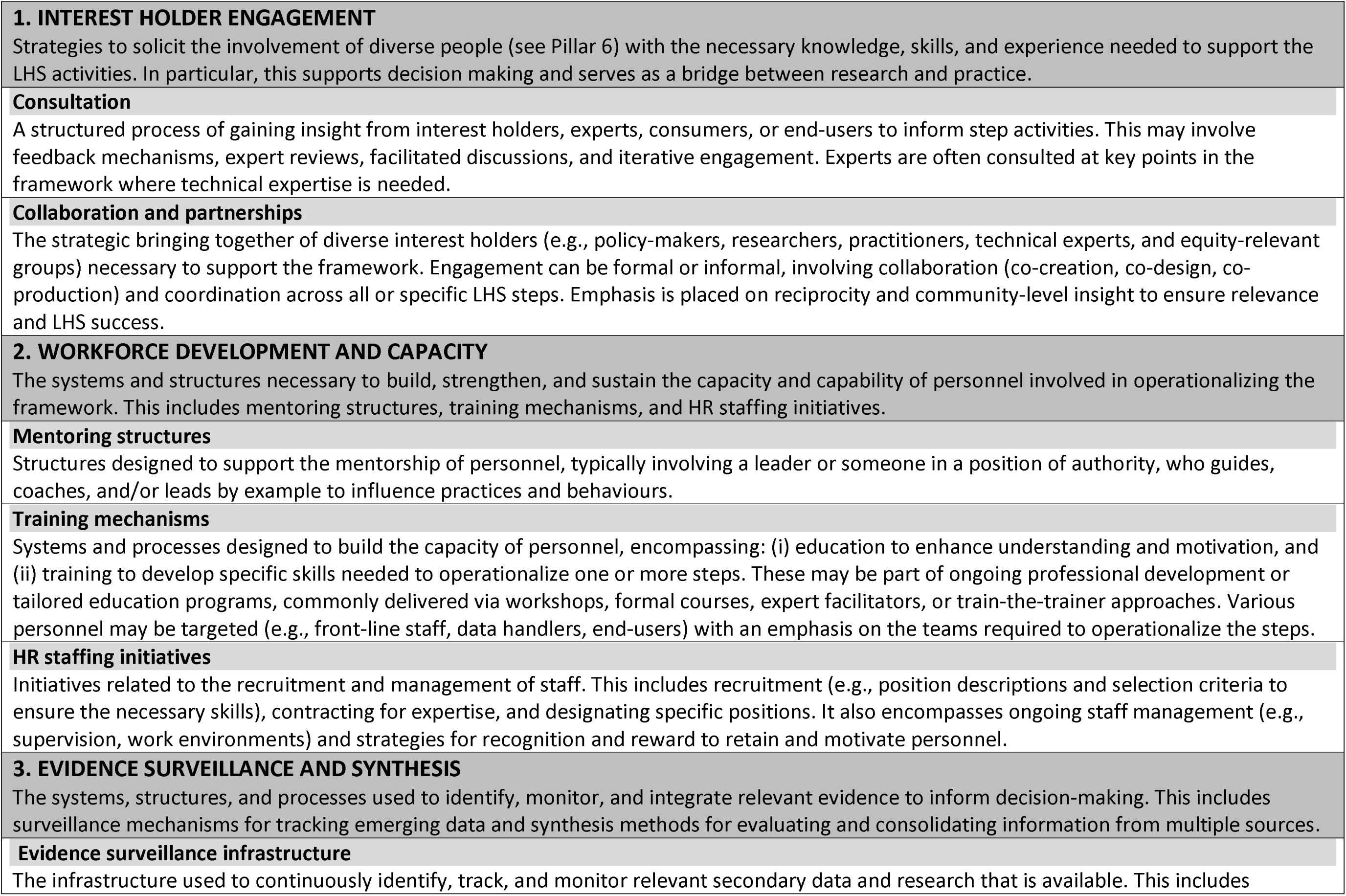
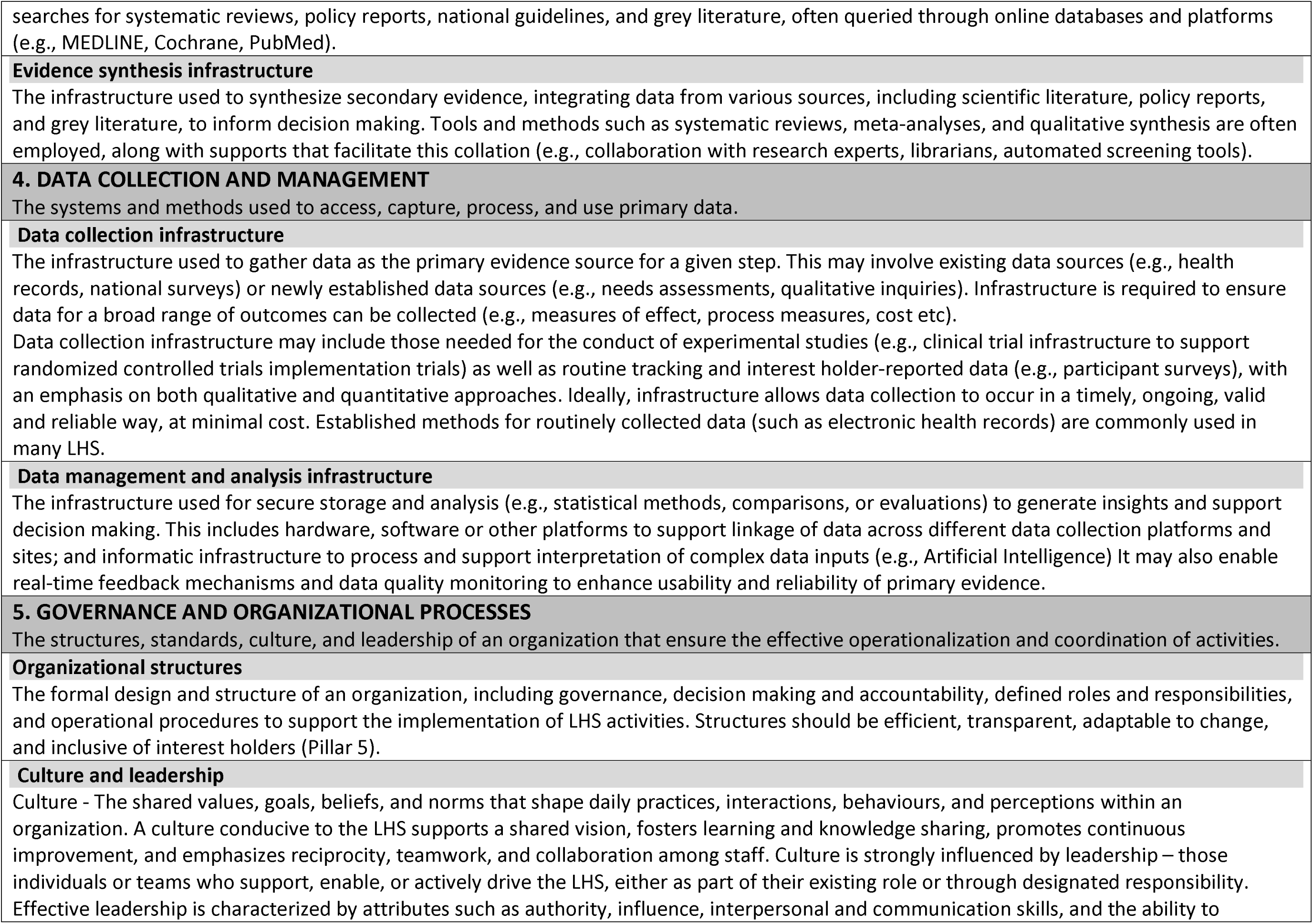
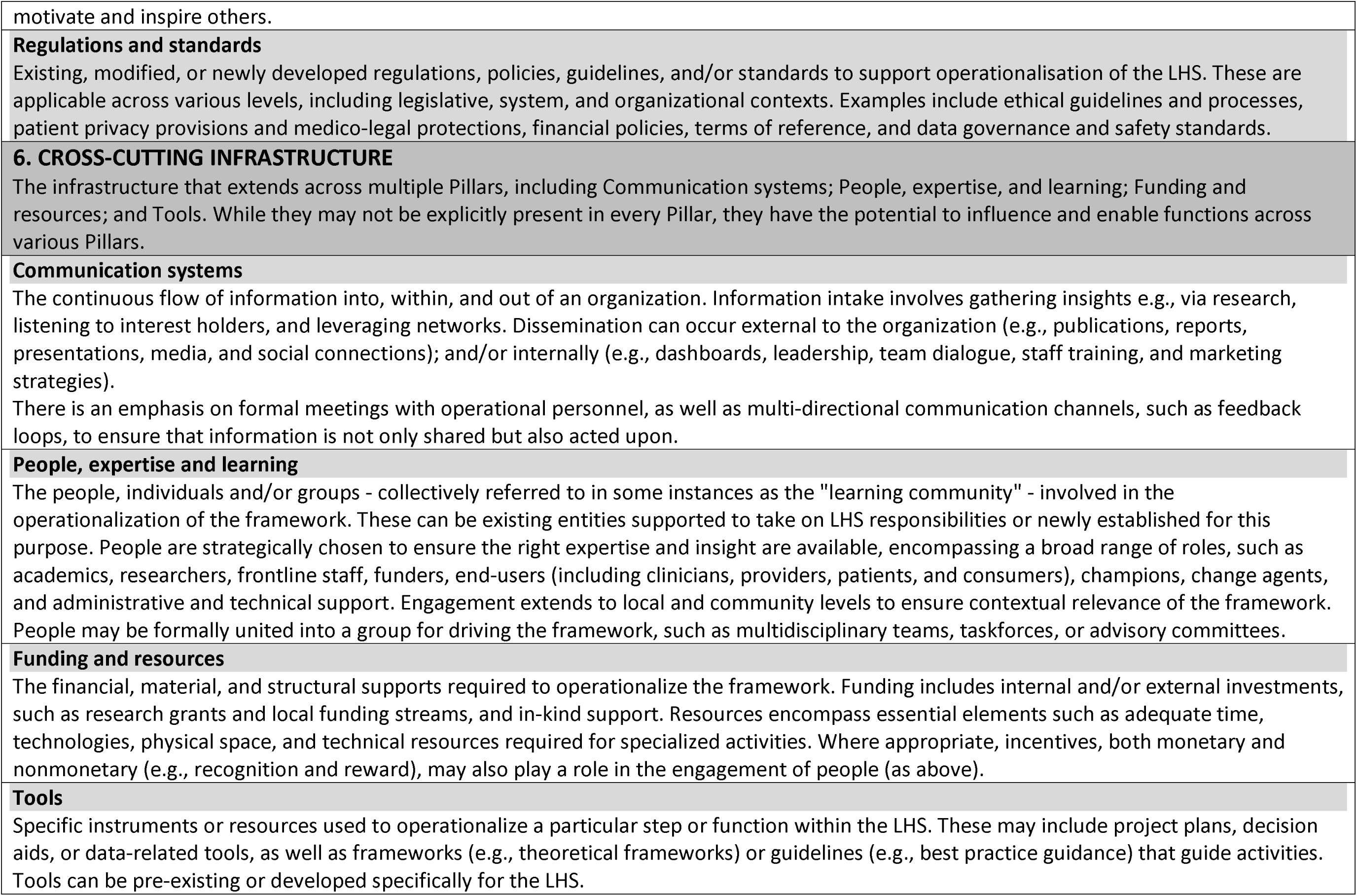
Pillars used to support a Learning Health System improvement cycle (steps)

### Objective 2: Steps

The steps data synthesis identified ten Steps across a three-phase LHS improvement cycle. Table 4 provides an overview of each Step and details of its core actions and activities. The coding presence for Steps across all guidance documents can be found in Supplementary File 5.

**Table 4.**
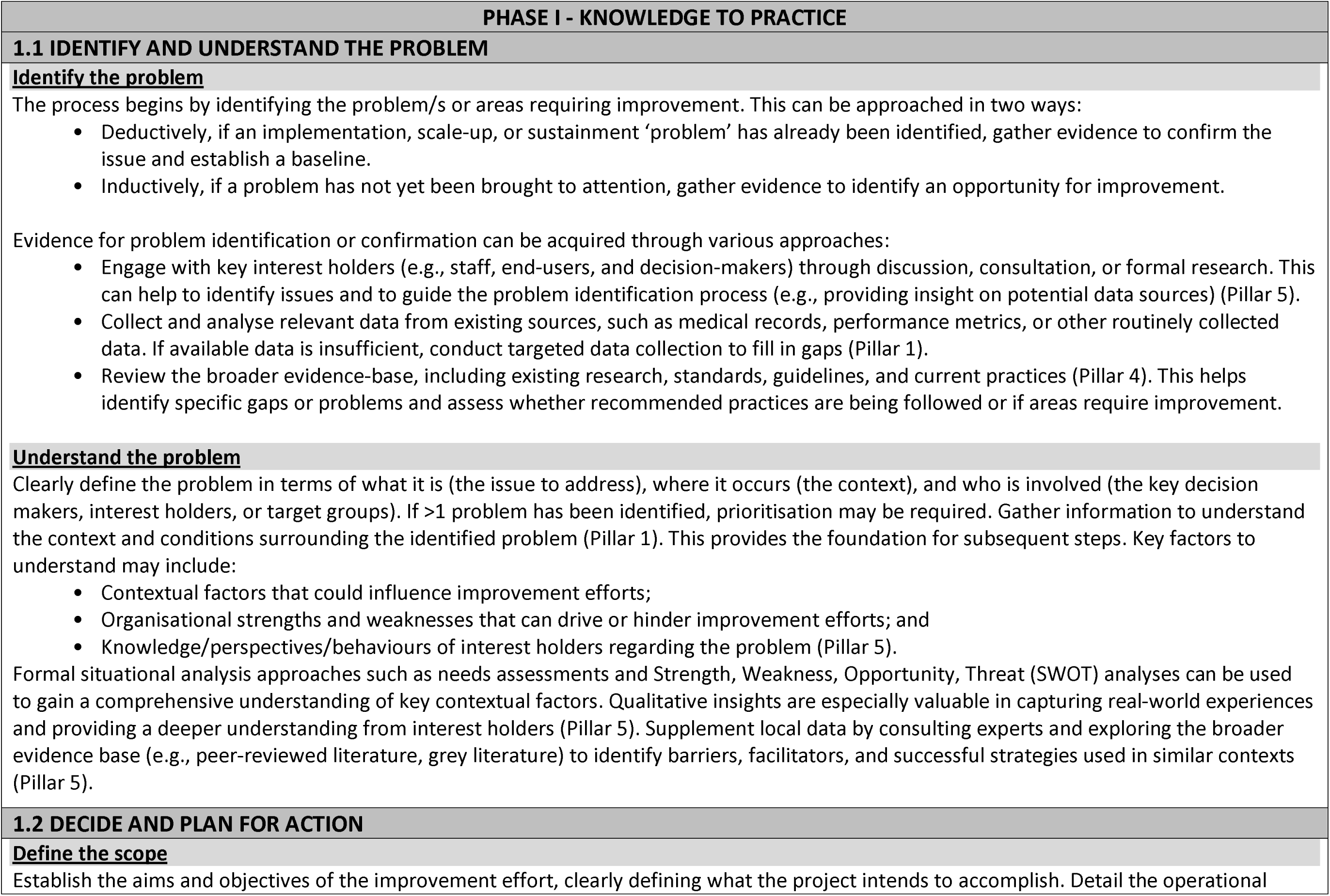
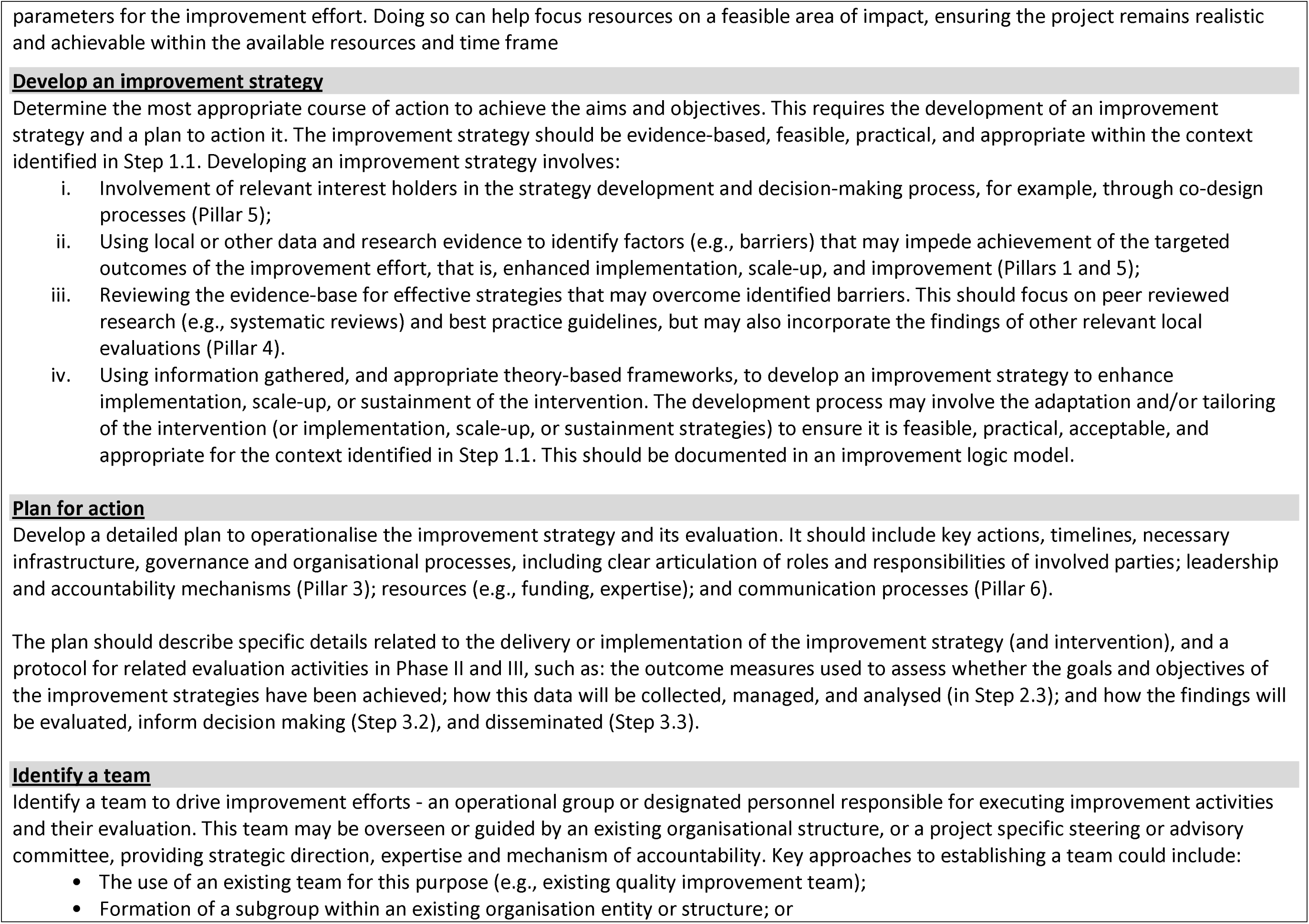
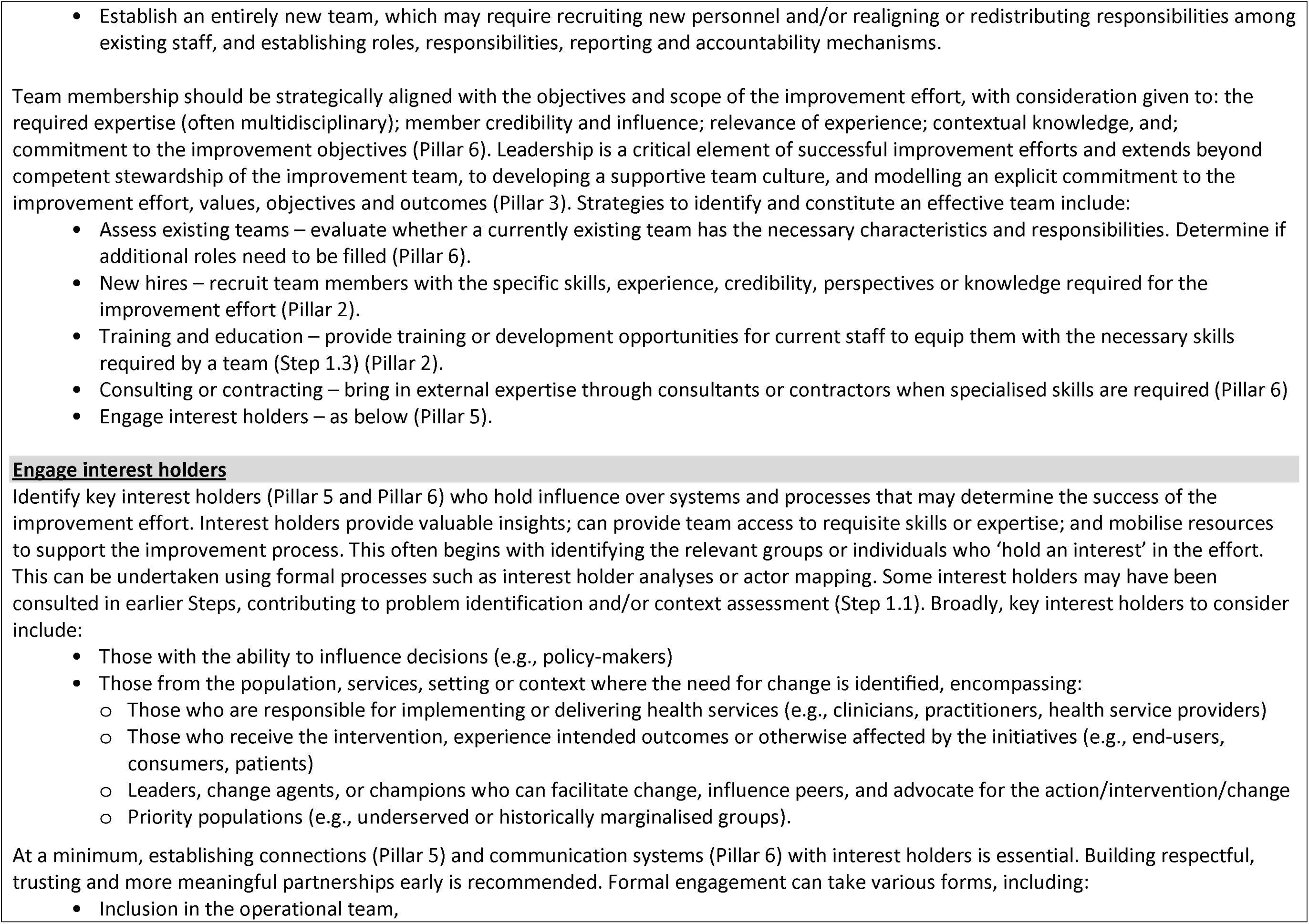
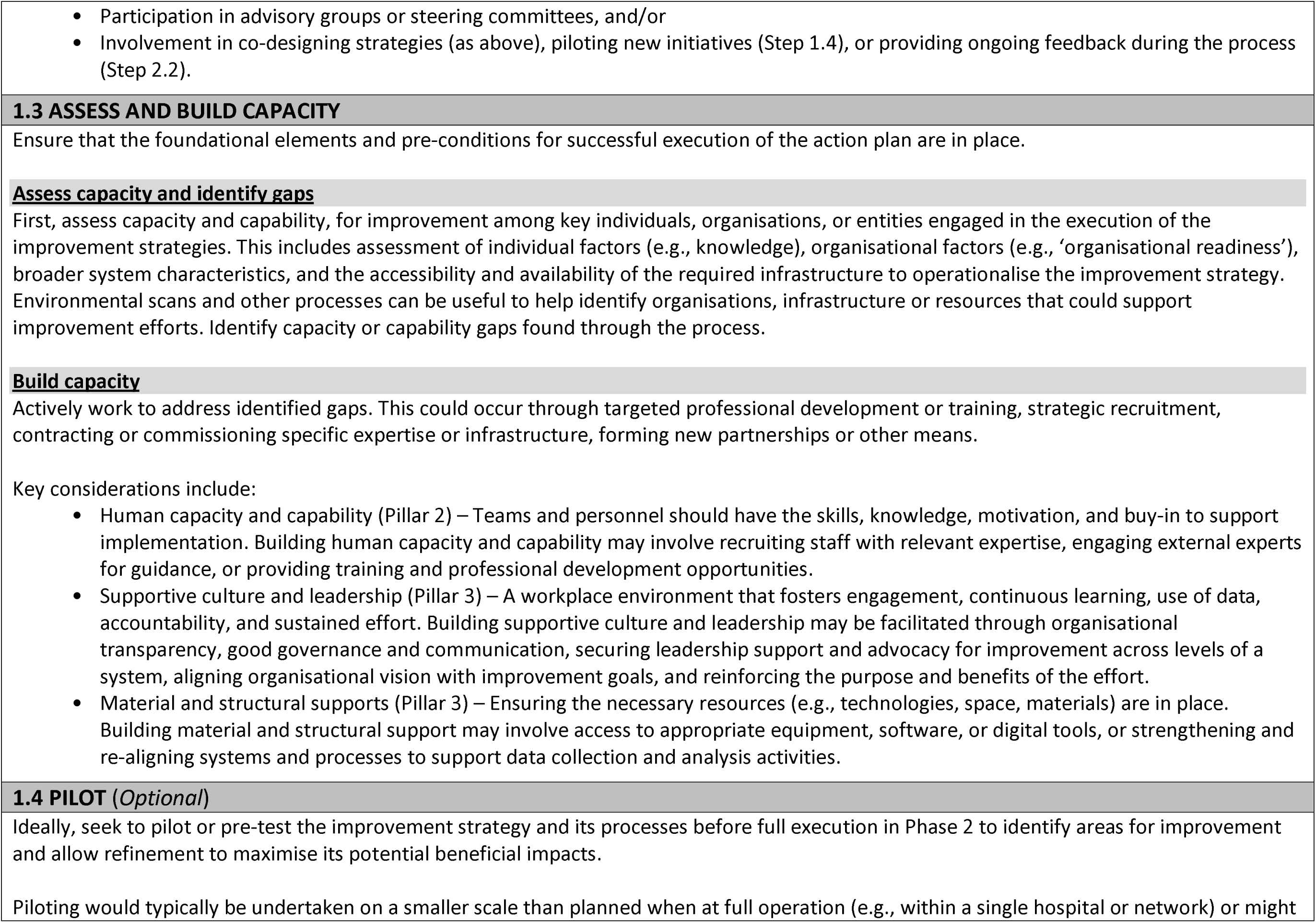
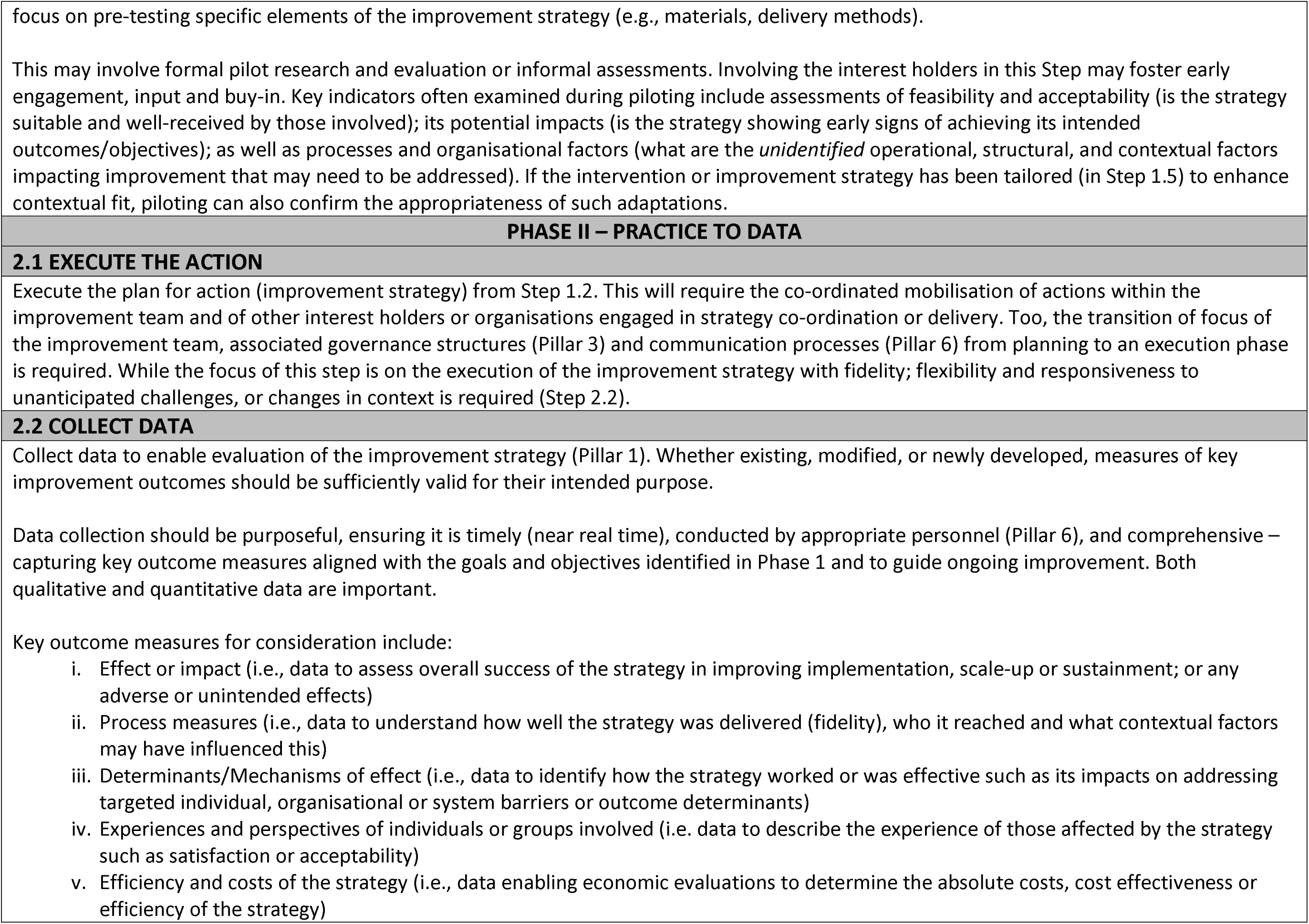
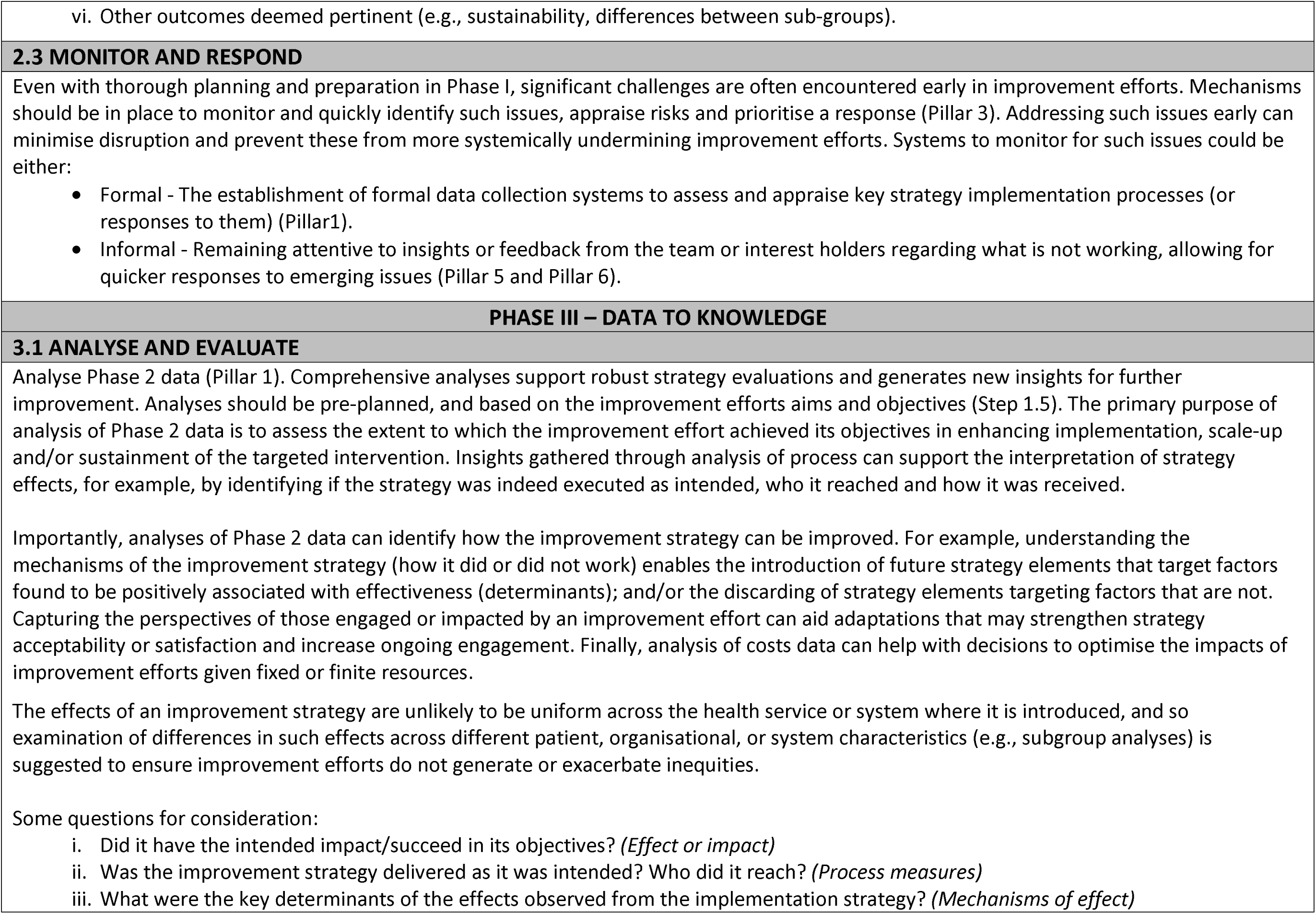
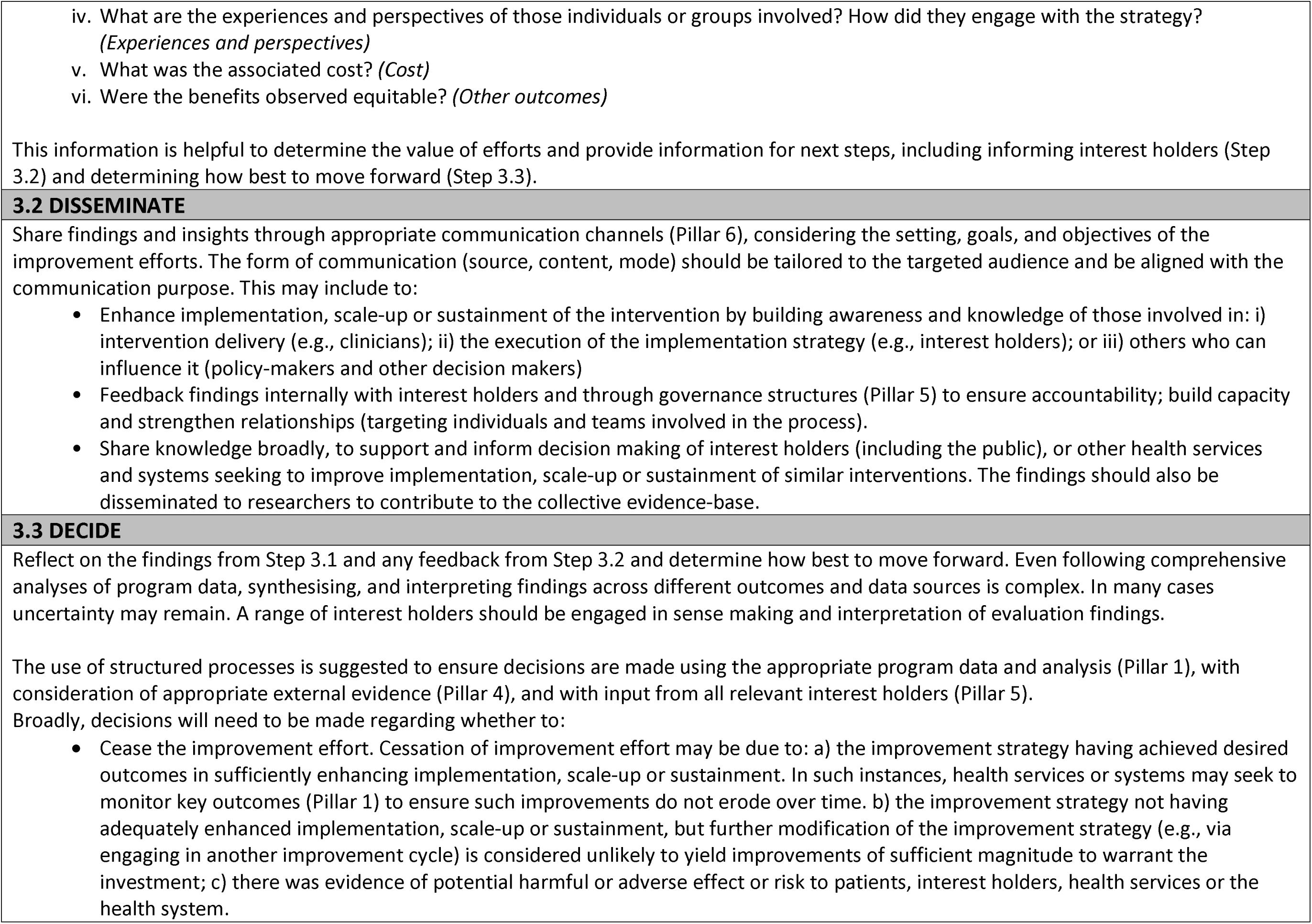
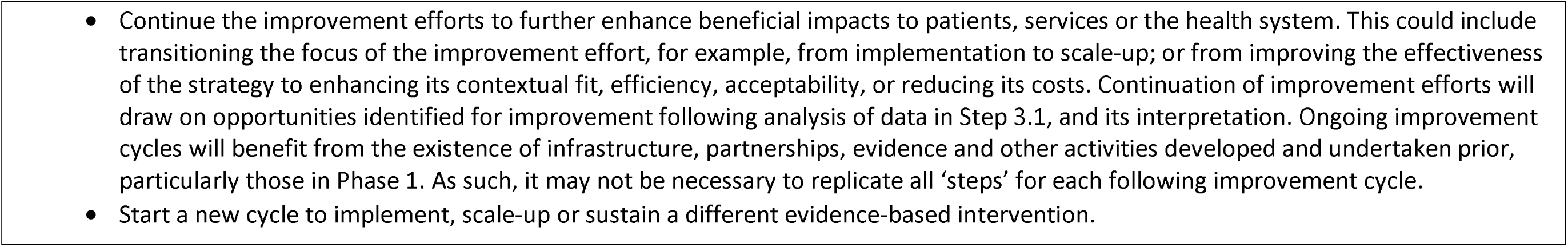
Steps used across a three-phase Learning Health System improvement cycle.

Phase 1) Knowledge to Practice, comprised four steps. Step 1.1 (Identify and understand the problem) involves the use of evidence to identify problem or areas requiring improvement, understanding its impacts and context. Step 1.2 (Decide and plan for action) includes developing an evidence-based strategy to address the identified problem; creating a detailed plan to operationalise the strategy and its evaluation, and identifying a team and interest holders to drive improvement efforts. Step 1.3 (Assess and build capacity) involves assessing capacity and capability to execute the action plan, and then actively working to address identified gaps – building capacity of the team, interest holders, and systems. Step 1.4 (Pilot) is an optional step, if feasible, and involves piloting or pre-testing the improvement strategy (activities to better implement, scale or sustain) and its processes before full execution, providing an opportunity to make refinements.

Phase 2) Practice to Data, comprised three steps. Step 2.1 (Execute the action) involves mobilising the team and supporting infrastructure to execute the action/improvement strategy. Step 2.2 (Collect data) involves collecting data to enable evaluation of the improvement strategy to assess or interpret its impacts. Step 2.3 (Monitor and respond) entails monitoring and quickly responding to any challenges or issues that are encountered when executing the improvement strategy.

Phase 3) Data to Knowledge, comprised three steps. Step 3.1 (Analyse and evaluate) involves analysing the collected data to assess the extent to which the improvement effort achieved its objectives. This data is used to identify opportunities to further improve the strategy. Step 3.2 (Disseminate) involves sharing the findings and insights internally or more broadly. Step 3.3 (Decide) involves reflecting on the findings and deciding how best to move forward – whether to cease, continue (by initiating another cycle), or shift the focus of the improvement effort.

### Sensitivity analyses

#### Quantitative coding comparison

Supplementary File 6 (Pillars) and 7 (Steps) display the results of the Crosstab Queries. As expected, slight variation in coding presence across Steps reflected the diverse nature and purpose of the guidance documents. For instance, some guidance focused heavily on early-phase planning (Phase 1; e.g., intervention design and preparation) while providing limited or no detail on subsequent phases. The highest coding presence was observed in Step 1.2 (Decide and plan for action; 99%), followed by Step 1.3 (Assess and build capacity; 58%). The lowest presence was for Step 2.3 (Monitor and respond; 14%) which was rarely identified in guidance documents as a distinct step. However, this activity was commonly noted in general recommendations or embedded within other steps, indicating its perceived importance despite limited explicit labelling.

When examined by framework characteristic, Step 1.1 (Identify and understand the problem) showed higher coding presence in local or organisational guidance (62%) compared to national or state-level guidance (27%). In other areas, meaningful comparisons were limited due to small sample sizes; for example, only seven guidance documents originated from low- and middle-income countries, and only three were intended primarily for academic audiences. Some coding absences were also conceptually appropriate; for example, frameworks focused on scale-up or sustainability often did not include Step 1.1, as these phases typically assume that the problem has already been identified.

For the Pillars, Pillar 6 (Cross-cutting infrastructure) had the highest coding presence, particularly for Communication systems (76%) and People and expertise (78%). Other highly represented Pillars included Pillar 5 (Governance and organisational processes; 83%) and Pillar 1 (Interest holder engagement; 70%). One observed difference was that Pillar 3 (Evidence surveillance and synthesis) was more commonly coded in national, or state-level guidance documents compared to local or organisational ones (82% vs 43%). Again, comparisons by framework characteristics were limited by small denominators.

Overall, the proportion of coded content for each Step and Pillar was relatively consistent across guidance documents with different characteristics. While some variability was observed, there were no notable outliers or dominant patterns. This suggests a balanced contribution from the diverse range of frameworks included in the review. This lends confidence that the final synthesis was not disproportionately influenced by any framework subtype and reflects a broad and inclusive representation of perspectives.

#### Qualitative coding comparison

The final Steps and Pillars were consistently applicable across all guidance subtypes, indicating strong coherence and relevance despite the diverse guidance documents. This suggests that we have captured a shared conceptual foundation underpinning guidance for improving implementation, scale-up, and sustainment of health interventions. While the overall structure of the Steps and Pillars was broadly transferable, the analysis also revealed nuanced differences in how they were applied or emphasised within certain subtypes of guidance. These distinctions were not observed between guidance documents by income setting (low- and middle-income country vs. high income country). Instead, distinctions were more evident across guidance subtypes, particularly based on the intended purpose of the guidance (e.g., scale-up or sustainability) or its policy orientation (e.g., national or state policy versus local or organisational policy).

For example, while the content of the Steps and Pillars was largely consistent across national or state policy and local or organisational policy guidance, a notable difference emerged in the sequencing of activities. In some local/organisational policy guidance documents, the process began with the identification or establishment of an operational team (e.g., Adapted Intervention Mapping [AIM] in schools^34^) to support implementation strategy selection and use in healthcare settings^35^, which then drove subsequent planning and implementation activities. This reflects one of the more challenging steps to place in a standardised sequence and is discussed further in the main discussion section.

Guidance documents focused specifically on scale-up often elaborated on or emphasised particular activities and structural considerations. These included the use of scalability assessments, distinguishing between different types of scale-up (e.g., vertical versus horizontal), and accounting for multiple organisational levels involved in scaling efforts (e.g., adopting units and scale-up entities). As a result, outcome measures and corresponding data systems (Pillar 1) were required to span both individual implementing units and system-wide levels. These documents also highlighted the broader implications of scale-up for personnel, underscoring the need for expanded interest holder engagement and enhanced team capacity (Pillars 5 and 6).

Similarly, guidance focused on sustainment placed greater emphasis on actions to assess and build infrastructure that could be supported over the long term (all Pillars). This included evaluating the availability of systems (data, training, communication etc.) and supports that could be maintained with existing resources of target organisations or settings. In particular, sustainment guidance emphasised the importance of robust data collection systems capable of ongoing monitoring (Pillar 1), highlighting that one-off assessments are typically insufficient for measuring sustainability outcomes.

Across both scale-up and sustainment guidance, working within and across resource-constrained settings was a common consideration. These documents frequently recommended strategies to optimise existing infrastructure, leverage partnerships, and build capacity using available resources. This reinforces the broader principle - reflected throughout our synthesis and elaborated in the Discussion section - that effective improvement activities are not solely about following prescribed steps but requires tailoring actions to context, including the realities of resource availability and system readiness.

## DISCUSSION

This comprehensive systematic review draws together LHS approaches to improvement with explicit synthesis of guidance to support the implementation, scale-up, and sustainment of evidence-informed health policies and interventions. We developed a consolidated guidance that comprises six pillars that provide the infrastructure, capacity, and expertise to support ongoing improvement activities. It also identified ten sequential yet flexible steps, organised across three LHS phases which comprised the continuous improvement process. It is hoped the work will contribute to the strengthening of health systems by supporting their generation and application of implementation (inclusive of scale-up and sustainment) research to more rapidly improve the safety and quality of healthcare, patient outcomes and community health.

A range of excellent LHS frameworks have been published^10–14^. These tend to focus on descriptions of the core infrastructure, or pillars needed to support improvement and ongoing learning. For example, Reid et al^11^ conceptualised an LHS as an engine with learning gears (such as implementation and evaluation) and factors such as leadership and funding providing the “fuel” needed for the gears to produce equity-centred outcomes. It also identifies several “moderators” and “brakes” that can restrict or even impede such outcomes. Similarly, Menear et al^12^ proposed a conceptual framework for value-creating LHSs that integrates four main dimensions – core values, pillars, processes, and outcomes. Their framework highlights seven pillars that support LHS functionality: scientific, social, technological, policy, legal, ethical, and socio-political infrastructure. These pillars are underpinned by core values such as transparency, equity, and collaboration, and are critical to enabling the learning processes that connect data to knowledge and action. Such ‘pillars’ provide an environment conducive to emergent learning, feedback and improvement within health systems. We saw value in consolidating such guidance and found remarkable consistencies in the attributes and functions of pillars across a diverse range of LHSs, and guidance documents. Such findings are similar to other recent reviews of LHS frameworks^10^. LHS frameworks provide less guidance, however, on the actual process of improvement - that is, the specific steps needed to enhance healthcare. Our review sought to address this gap by synthesising the steps recommended by process models of three important and distinct outcomes that help maximise the impact of evidence-based healthcare: implementation, scale-up, and sustainment. It is often suggested that each of these outcomes are considered across each phase of an improvement process. For example, sustainment should be considered when selecting an intervention to implement, and in the process of implementation. We are not aware, however, of guidance that has attempted to bring these together. While the application of the process may differ, such as the specific barriers, strategies employed to address them, or measures used to assess outcomes, we found little differences in the broad steps or process reported in documents reviewed between those with an emphasis on implementation, scale-up or sustainment. We believe this is useful contribution, offering a structured, but consistent approach to improvement. Importantly, the process is also not intended to be static. In alignment with the continuous nature of LHS, it can be re-applied over time to support different stages of improvement – from initial implementation through to scale up or sustainment. In this way, it offers both common process and practical flexibility for strengthening evidence-informed practice in dynamic, real-world settings.

Our findings closely align with the theoretical domains of evidence-informed policy-making (EIP) institutionalisation described by Kuchenmuller et al^36^. Their framework identifies six domains essential for embedding evidence use within policy systems: governance, standards and routinised processes, partnership, collective action and support, leadership and commitment; resources, and culture. These domains map closely onto our identified Pillars. However, while the EIP institutionalisation framework primarily conceptualises these domains as structural and cultural mechanisms that confer legitimacy and sustain evidence use, our synthesis focuses on their practical operationalisation within LHSs, specifically in relation to implementation, scale-up, and sustainment of health interventions. In particular, our findings place emphasis on the systems and processes needed to generate new practice-based evidence, with dedicated Pillars for data systems and evidence synthesis. Such elements are not foregrounded in Kuchenmüller et al.’s framework^36^, which assumes the availability of existing evidence for use. This focus on evidence generation, as well as use, underscores the dual role of LHSs as both users and producers of evidence. Additionally, while Kuchenmüller et al. frame institutionalisation as a process unfolding in stages, our findings describe a cyclical, practice-oriented improvement process, comprising actionable steps. These two perspectives - the institutionalisation of evidence use and the operationalisation of implementation improvement cycles - offer complementary pathways toward strengthening LHSs. Future research could explore how these perspectives can be integrated to support durable, evidence-informed health systems globally.

Synthesising the steps or stages across guidance documents was a complex process. Unlike the pillars, which do not follow a prescribed order, the steps were typically presented as sequential activities with variation in how different guidance documents presented this sequence. Some did not present any explicit ordering at all. Despite this challenge, we were able to generate a coherent and comprehensive set of steps through iterative coding processes and remaining grounded in the overall objectives and intended users of the review. In cases where ambiguity existed, we prioritised placement that aligned with the practical needs of policy-makers and targeted end users. To enhance usability and relevance across settings, we deliberately kept the steps broad. In some cases, this meant including several key actions (i.e., sub-steps) within a single step, to better reflect the fluid and often non-linear nature of real-world improvement processes.

The Phase 1 steps were especially challenging to position. For example, the identification and establishment of an operational team appeared at various points across guidance documents. In some cases, it was presented more as a pillar – a resource either assumed to be already present or meant to be established outside of the guidance application. For example, the Competitive Intelligence Process described a network of people (including executives and those with designated positions) as an infrastructure to operationalise the process, but not as step within the process itself ^37^. Another challenge was interest holder identification and engagement – this was a recurring activity across all phases of the guidance. As such, we addressed this aspect by including it within Step 1.2 as well as within one of the six pillars, recognising its importance as an infrastructure across the entire improvement process. We also included the key interest holder-related information and activities within respective steps – for example, engaging interest holders to help identify and understand the problem (Step 1.1), and the formal identification of interest holders needed for improvement activities as part of planning in Step 1.2. Despite challenges in synthesis, the overarching LHS phases^29^ provided a highly useful structure for organising the steps. These phases were unchanged from the outset of synthesis, offering a strong scaffold for coding and situating the diverse improvement activities described across the guidance documents.

A number of methodological strengths of the review are worth noting. The coding process was primarily inductive, allowing for findings to emerge directly from the source data, supported by strong LHS-based codebooks and a collaborative coding team. Where gaps or inconsistencies arose, deductive searches and peer discussions were used to clarify concepts and validate emerging interpretations. This experience highlights the value in having content expertise within the coding team and involved across the synthesis process, to identify potential gaps and ensure thorough coding. All coded data were reviewed by at least three members of the research team: original coders, secondary reviewers, and a third reviewer during the final data cleaning process. This ensured a high level of consistency and rigor.

Unlike traditional reviews and syntheses that appraise the quality of included documents to determine the strength of the evidence base, our approach treated all included guidance documents equally. Given the purpose of this review, we did not apply differential weighting based on document quality or comprehensiveness. Instead, all documents were reviewed in full, and all relevant content was extracted, regardless of whether a document provided a comprehensive framework or focused on a single component (e.g., strategy development or evaluation). While some documents contributed more coded content than others, this did not lead to greater influence on the results. The final steps and pillars were formed using an iterative, team-based synthesis process, using the breadth of relevant guidance across all included documents.

However, the comprehensive nature of steps and pillars also presents a potential limitation. At first glance, the steps and pillars may appear complex or even infeasible, particularly for settings with limited resources or capacity. Yet this reflects its purpose: to present a thorough version of what LHSs focused on implementation, scale-up, and sustainment can entail. Importantly, the review findings do not assume that all components must be enacted in full, and while the ‘steps’ are presented linearly, in complex adaptive systems like healthcare, we acknowledge this will rarely be the case. The application of findings from this review will be different by setting and context; the steps and pillars should be interpreted and tailored to the context in which they are applied. This tension between ideal and feasible is not unique to our synthesis - most individual guidance documents included in this review explicitly acknowledged the challenges posed by contextual constraints and resourcing limitations. For example, the IDEAS (Integrate, Design, Assess, and Share) framework drew attention to the variability of its application based on resource constraints ^38^.

Nonetheless, there are likely to be challenges in operationalising comprehensive LHSs consistent with the steps and pillars describe in this review. ^39^. Health systems operate within complex, resource-constrained environments, and establishing and sustaining multi-disciplinary teams, building the necessary infrastructure, and embedding continuous learning cycles into routine practice can be resource- and time-intensive. It may also require shifts in culture, processes, and roles. Many of the included guidance documents acknowledged the importance of a supportive organisational culture in enabling reorientation for improvement efforts. For example, the LHS Architectural Framework emphasised the need for internal alignment and a culture of inquiry to embed continuous learning into routine operations^40^. For academics and public health researchers, it also represents a new way of working^29^, one that requires closer, ongoing partnerships and a willingness to adapt timelines and traditional processes. Potential risks include insufficient leadership buy-in, fragmented data systems, or a lack of resourcing to enact the full cycle. The potential for health systems to improve the quality and impact of healthcare is unrealised. There are a range of challenges to developing learning health systems, even in high income countries and well-resourced healthcare organisations. This review sought to facilitate the development of LHSs by consolidating diverse guidance, to enhance the generation and application of research to support better implementation, scale-up and sustainment of effective health interventions. Here we provide key findings of our synthesis. The WHO will soon publish a practical guide to support to help operationalise LHSs. Further efforts should focus on testing and refining the framework in real world contexts to further strengthen its relevance and impact.

## Supporting information

Supplementary File 1. Search Strategy

Supplementary File 2. Website Searches

Supplementary File 3. Characteristics of included documents

Supplementary File 4. Pillars coding per document

Supplementary File 5. Steps coding per document

Supplementary File 6. Pillars quantitative comparison

Supplementary File 7. Steps quantitative comparison

## Data Availability

All data produced in the present study are available upon reasonable request to the authors

## ACKNOWLEDGEMENTS

The authors would like to acknowledge the support of Megan Duffy, Kate Bartlem, Rebecca Hodder, Bronwen Merner, and Nicole Nathan for their support with the initial stages of this study. We also thank the members of the WHO editorial board convened for the development of the guidance on implementation, scale-up, and sustainment of evidence-informed health interventions.

## SUPPORTING INFORMATION CAPTIONS

S1 **Supplementary File. Search strategy**

**S2 Supplementary File. Website searches**

**S3 Supplementary File. Characteristics of included documents**

**S4 Supplementary File. Pillars coding per document**

**S5 Supplementary File. Steps coding per document**

**S6 Supplementary File. Pillars quantitative comparison**

**S7 Supplementary File. Steps quantitative comparison**

## DECLARATIONS

### Contributions

LW conceived of this study and led the initial conceptual planning. CL and SMc supported development of the methodology. SMc, HT, LC, MW, CB, SG and MF conducted screening and/or data extraction. CL led evidence syntheses. CL, SMc, HT, MW, SG, CB, LC and LW coded data and contributed to the development of pillars and steps. All authors were involved in the interpretation of the results. CL and SMc drafted the manuscript, and all authors contributed to and approved the final version. The corresponding author attests that all listed authors meet authorship criteria and that no others meeting the criteria have been omitted.

The authors alone are responsible for the views expressed in this article and they do not necessarily represent the views, decisions or policies of the institutions with which they are affiliated.

### Data sharing

Data is available to be shared upon reasonable request.

### Ethical approval

Ethical approval was not required to be obtained for this study.

