## Supplementary File 1. Search Strategy for "Learning Health System for implementation, scale-up, and sustainment: A systematic review to consolidate guidance for improvement"

Medline (OVID) search date 12^th^ April 2024

| **#** | **Searches** |  |
| --- | --- | --- |
| 1. | implement*.mp. | 751071 |
| 2. | adopt*.mp. | 377696 |
| 3. | (system* adj2 change*).mp. | 21380 |
| 4. | quality improvement*.mp. | 75409 |
| 5. | sustain*.mp. | 504778 |
| 6. | institutionali*.mp. | 23038 |
| 7. | routin*.mp. | 503992 |
| 8. | scale*.mp. | 1237543 |
| 9. | “learning health* system*” | 1341 |
| 10. | (guideline* OR guidance OR (practice ADJ2 (guide*1 OR recommend* OR standard*)) OR (decision* ADJ2 (making OR make*)) OR (evidence-based ADJ2 (practice* OR medicine OR nursing))).ti,ab. | 925280 |
| 11. | (or/1-9) adj5 10 | 41619 |
| 12. | (Health care or healthcare).ti,ab. | 769065 |
| 13. | Clinical practice.ti, ab. | 265414 |
| 14. | Health service*.ti,ab. | 140773 |
| 15. | Public health.ti,ab. | 345189 |
| 16. | Community health.ti,ab. | 31158 |
| 17. | Community service.ti,ab. | 1995 |
| 18. | Medical Care.ti,ab. | 62880 |
| 19. | Health polic*.ti,ab. | 36589 |
| 20. | or/12-19 | 1462241 |
| 21. | 11 and 20 | 15018 |
| 22. | Randomized Controlled Trial/ | 610363 |
| 23. | clinical trial/ or controlled clinical trial/ | 564058 |
| 24. | random allocation/ | 107073 |
| 25. | Double-Blind Method/ | 177958 |
| 26. | Single-Blind Method/ | 33354 |
| 27. | placebos/ | 35934 |
| 28. | Research Design/ | 1504529 |
| 29. | Evaluation Studies/ | 127580 |
| 30. | Comparative Study/ | 262079 |
| 31. | exp Longitudinal Studies/ | 1913805 |
| 32. | exp Longitudinal Studies/ | 170646 |
| 33. | exp Cohort studies/ | 56433 |
| 34. | Controlled Before-After Studies/ | 2590910 |
| 35. | Interrupted Time Series Analysis/ | 752 |
| 36. | comparative study.pt. | 2019 |
| 37. | clinical trial.tw. | 1913805 |
| 38. | latin square.tw. | 201638 |
| 39. | (time adj series).tw. | 5549 |
| 40. | (before adj2 after adj3 (stud* or trial* or design*)).tw. | 48137 |
| 41. | ((singl* or doubl* or trebl* or tripl*) adj5 (blind* or mark)).tw. | 17351 |
| 42. | placebo*.tw. | 202356 |
| 43. | random*.tw. | 255103 |
| 44. | (matched adj (communit* or school* or population*)).tw. | 3335 |
| 45. | control*.tw. | 4784578 |
| 46. | (comparison group* or control group*).tw. | 616421 |
| 47. | matched pairs.tw. | 8124 |
| 48. | outcome stud*.tw. | 9437 |
| 49. | (quasiexperimental or quasi experimental or pseudo experimental).tw. | 22258 |
| 50. | (nonrandomi?ed or non randomi?ed or psuedo randomi?ed or quasi randomi?ed).tw. | 40636 |
| 51. | prospectiv*.tw. | 911314 |
| 52. | volunteer*.tw. | 221662 |
| 53. | (or/22-52) | 9683503 |
| 54. | 21 not 53 | 9912 |
| 55. | limit 54 to (last 10 years) | 6853 |
