## Supplementary File 2. Website Searches for "Learning Health System for implementation, scale-up, and sustainment: A systematic review to consolidate guidance for improvement"

### Supplementary website searches

<https://www.nccmt.ca/organizational-change>

<https://www.ahrq.gov/>

<https://thecenterforimplementation.com/toolbox>

<https://impsciuw.org/>

<https://aho.afro.who.int/af>

<https://campus.paho.org/en>

<https://www.paho.org/en/technical-and-scientific-products>

<https://www.who.int/southeastasia/activities>

<https://www.emro.who.int/e-library/index.html>

<https://www.who.int/westernpacific/our-work/resources>

<https://www.who.int/europe/about-us/our-work>

<https://evidence-impact.org/>

<https://www.emro.who.int/evidence-data-to-policy/training-package/index.html>
